# The social determinants associated with the advanced stage diagnosis of breast cancer in Egypt

**DOI:** 10.1101/2023.08.10.23293931

**Authors:** Madonna A. Fekry, Loay Kassem, Justin C. Yang, Hamdy A. Azim

**Affiliations:** Clinical Research Department, Cairo Oncology Center, Cairo, Egypt; Clinical Oncology Department, Kasr Alainy School of Medicine, Cairo University, Cairo, Egypt; Medical Oncology Unit, Cairo Oncology Center, Cairo, Egypt; Division of Psychiatry, University College London, United Kingdom; Camden & Islington NHS Foundation Trust, United Kingdom; Centre for Public Health, Wolfson Institute of Population Health, Queen Mary University of London, United Kingdom

## Abstract

**Background:** Majority of Egyptian breast cancer (BC) patients present at advanced stages. We examined the sociodemographic and clinical factors associated with late presentation of BC.

**Methods:** This is a retrospective cohort study of patients who presented with BC between 2011 and 2020. Logistic regression was performed to examine the association between sociodemographic factors and advanced BC.

**Results:** This cohort included 1,953 patients with median age of 52 years. 1,098 (56.2%) patients were diagnosed at early stages, while 855 (43.8%) patients were diagnosed at advanced stages. Univariate logistic regression analyses revealed that several sociodemographic and clinical factors were associated with advanced BC, including having negative family history of BC (OR= 0.1.27; 95% CI:1.05 – 1.54), having no job (OR= 1.28; 95% CI: 1.06 – 1.53), being married (OR=1.27; 95% CI:1.02 – 1.57), rural residence (OR=1.27; 95% CI:1.02 – 1.57), having more than three children (OR=1.42; 95% CI:1.15 – 1.75), higher KI-67% score (OR=1.01; 95% CI:1.00 - 1.01), having HER2-Enriched or TNBC subtypes (OR= 1.44; 95% CI:1.16 – 1.79), and having tumor grate II/III (OR=4.12; 95% CI:1.58 – 10.77 & OR= 1.44; 95% CI:1.16 – 1.79, respectively). In the multivariate logistic regression model, only KI-67% (aOR=1.01; 95% CI:1.00 - 1.02), having no job (aOR=1.44; 95% CI:1.10 – 1.90), and rural residence (aOR=1.88; 95% CI:1.03 – 3.42) were significantly associated with advanced BC.

**Conclusion:** This study concluded that having no job and rural residence are highly associated with advanced BC. Raising the public awareness is the best strategy to encourage early detection of BC.

## Background

Breast cancer (BC) is the most common cancer and the second leading cause of cancer death among women worldwide. In 2020, there were 2.3 million new breast cancer cases and 685,500 breast cancer related deaths globally (1). In Egypt, there were 22,038 new BC cases and 9,148 BC related deaths in 2020 (2). Nonetheless, according to a recent meta-analysis conducted by our group, 56% of Egyptian BC patients were diagnosed at stages III-IV and 70% presented with node-positive disease (3). Despite the obvious dominance of late disease presentation, little is known about the sociodemographic determinants of BC diagnosis at advanced stages in Egypt.

Early-stage BC is a curable disease. Conversely, advanced stage BC is associated with significantly inferior survival (4). In July 2019, the Egyptian Women’s Health Initiative was launched. The initiative aimed at reducing the frequency of late-stage disease to below 30% by July 2025 via a national screening program using novel 2-step clinical examination platforms. There was an excellent uptake of the early detection program with more than 20,000,000 women screened so far (5). Even though a shift towards downstaging strategy is important, it cannot be achieved by only establishing early detection programs. Observing the sociodemographic factors that are associated with rate of breast cancer screening performance and BC stage at diagnosis is as important as increasing population awareness of breast cancer and its screening techniques. Globally, sociodemographic disparities, such as education, race, and socioeconomic status are associated with advanced stage diagnosis of BC (6).

Nevertheless, there is limited evidence on the association between such determinants and BC stage at diagnosis in Egypt. Therefore, the aim of this study is to examine the sociodemographic and clinical factors associated with late presentation and detection of BC in Egypt. Accordingly, this can guide policy adjustments towards better outcomes.

## Methods

### Study design and data collection

This retrospective cohort study included 1,953 patients with BC who presented to Cairo Oncology Center (COC) between 2011 and 2020. The study was approved by the local IRB in November 2022. Data were extracted retrospectively from the patients’ medical files from October to November 2022. The variables that have been collected are sociodemographic variables (e.g., city of residence, employment status, religion, number of children, and marital status), clinical characteristics (e.g., height, weight, age at diagnosis, gender, family history of BC, and menopausal status), and pathological characteristics (e.g., histopathological type, tumor grade, tumor size, T stage, N stage, M stage, ER status, PR status, HER-2 status, KI-67 status, and status at first presentation). The outcome variable analyzed in this cohort was advanced stage diagnosis of BC. The 7th edition of the American Joint Committee on Cancer (AJCC) staging system for BC was used in this cohort. Early-stage BC was defined as stage I and stage II, while advanced stage BC was defined as stage III and stage IV.

### Statistical analysis

SPSS (version 20.0; IBM Corp, Armonk, NY), JASP (version 0.16.1.0), R Program (version R-3.6.1), and Tableau (version 2021.3) were used for running the whole data analysis and visualization. Furthermore, 2-sided p value <.05 was considered statistically significant. Chi-square test or Fisher’s exact test were used to examine the association between stage at diagnosis and different variables (sociodemographic and biological variables). Shapiro Wilk test and Test of Equality of Variances were used to check on the normality and homogeneity assumptions of the continuous numeric variables. Independent Samples T-test or Wilcoxon rank-sum test were used to examine the mean difference in continuous variables (age, BMI, and KI-67% score) between patients who presented at early stages and those who presented at advanced stages. Univariate logistic regression analysis was used to examine the odds of presenting with advanced BC. Variables that showed a significant association of p<0.05 were included in the multivariate logistic regression analyses.

## Results

### Baseline characteristics of the patients’ cohort

The cohort included 1953 BC patients with median age of 52 years (IQR: 43 – 61). The cohort consisted of 15 (0.8%) males and 1,939 (99.2%) females. Among the 1,939 females, 992 (55.6%) were postmenopausal and 792 (44.4%) were premenopausal. 1,098 (56.2%) patients were diagnosed at early stages, while 855 (43.8%) patients were diagnosed at advanced stages. Patients who aged 40 years or less constituted 20% of the cohort, while patients who aged more than 40 years constituted 80%. The median body mass index (BMI) of the whole cohort was 30.8 (IQR: 26.7 – 35.1). The median KI-67% score of the whole cohort was 20 (IQR: 10 – 30). 1343 (68.7%) patients had negative family history of BC, while 611 (31.3%) patients had positive family history of BC. 1080 (56.5%) patients had no job, while 831 (43.5%) patients had paid job. At diagnosis time, 457 (23.9%) were unmarried and 1454 (76.1%) were married. 128 (6.8%) patients were rural residents, while 1757 (93.2%) were urban residents. For the city of residency, 1214 (63.3%) patients resided in Cairo and Giza and 703 (36.7%) patients resided in other 22 cities in Egypt. 111 (5.8%) patients were Christians, while 1803 (94.2%) were Muslims. At presentation time, 1378 (74.8%) patients had three children or fewer and 465 (25.2%) patients had more than three children. 452 (25.8%) patients had either HER2-Enriched or TNBC subtypes, while 1301 (87.7%) patients had luminal A/B-like subtypes. The median KI-67% score of the whole cohort was 20 (IQR: 10 – 30). For the tumor grade, 32 (1.9%) patients had grade I, 1400 (84.6%) had grade II, and 222 (13.4%) had grade III. Table 1 shows the baseline characteristics of the whole cohort.

**Table 1.** Characteristics of patients diagnosed at early stage versus advanced or metastatic stage.

|  | Early stage<br>n= 1098 | Advanced or<br>metastatic stage<br>n= 855 | Total<br>n= 1953 | p-value |
| --- | --- | --- | --- | --- |
| Sociodemographic variables |  |  |  |  |
| Age (years) (mean ± SD) | 52.3 ± 12.43 | 51.23 ± 12.29 | 51.83 ± 12.38 | 0.056 |
| Age categories |  |  |  |  |
| 40 years or less | 210 (19.1%) | 181 (21.2%) | 391 (20.0%) | 0.263 |
| More then 40 years | 888 (80.9%) | 674 (78.8%) | 1562 (80.0%) |  |
| Body mass index (kg/m2) (mean ± SD) | 31.08 ± 6.69 | 31.66 ± 7.24 | 31.34 ± 6.93 | 0.219 |
| KI-67% score (mean ± SD) | 23.87 ± 21.06 | 26.65 ± 20.31*** | 24.91 ± 20.82 | <0.001 |
| Family history of breast cancer |  |  |  |  |
| Negative family history | 730 (66.5%) | 612 (71.6%) | 1343 (68.7%) | 0.016 |
| Positive family history | 368 (33.5%)* | 243 (28.4%) | 611 (31.3%) |  |
| Employment status |  |  |  |  |
| No job (housewife, retired, or student) | 579 (53.9%) | 501 (59.9%)** | 1080 (56.5%) | 0.009 |
| In paid job | 495 (46.1%) | 336 (40.1%) | 831 (43.5%) |  |
| Marital status |  |  |  |  |
| Unmarried (single, divorced, or widow/widower) | 277 (25.8%)* | 180 (21.5%) | 457 (23.9%) | 0.031 |
| Married | 798 (74.2%) | 656 (78.5%) | 1454 (76.1%) |  |
| Urbanicity |  |  |  |  |
| Rural | 53 (5.0%) | 75 (9.1%)** | 128 (6.8%) | 0.001 |
| Urban | 1005 (95.0%) | 752 (90.9%) | 1757 (93.2%) |  |
| City of residence |  |  |  |  |
| Big cities (Cairo and Giza) | 708 (66.0%) | 506 (60.0%) | 1214 (63.3%) | 0.012 |
| Other 22 cities | 365 (34.0%) | 338 (40.0%)** | 703 (36.7%) |  |
| Religion |  |  |  |  |
| Christian | 58 (5.4%) | 53 (6.3%) | 111 (5.8%) | 0.398 |
| Muslim | 1016 (94.6%) | 787 (93.7%) | 1803 (94.2%) |  |
| Parity |  |  |  |  |
| 3 or fewer children | 806 (77.6%) | 572 (71.1%) | 1378 (74.8%) | 0.001 |
| More than 3 children | 232 (22.4%) | **233 (28.9%)\*\*** | 465 (25.2%) |  |
| Gender |  |  |  |  |
| Male | 6 (0.5%) | 9 (1.1%) | 15 (0.8%) | 0.204 |
| Female | 1092 (99.5%) | 846 (98.9%) | 1939 (99.2%) |  |
| **Biological variables** |  |  |  |  |
| Breast cancer subtypes |  |  |  |  |
| HER2E or TNBC | 234 (23.3%) | **218 (29.2%)\*\*** | 452 (25.8%) | 0.005 |
| Other subtypes (Luminal A, B1 and B2) | 772 (76.7%) | 529 (70.8%) | 1301 (87.7%) |  |
| Tumor grade |  |  |  |  |
| Grade I | **27 (2.9%)\*\*** | 5 (0.7%) | 32 (1.9%) | 0.007 |
| Grade II | 794 (84.1%) | 606 (85.4%) | 1400 (84.6%) |  |
| Grade III | 123 (13%) | 99 (13.9%) | 222 (13.4%) |  |
| Menopausal status |  |  |  |  |
| Postmenopausal | 574 (56.9%) | 418 (53.9%) | 992 (55.6%) | 0.213 |
| Premenopausal | 435 (43.1%) | 357 (46.1%) | 792 (44.4%) |  |
| Bold indicates statistically significant findings by Chi-square and Wilcoxon rank-sum tests |  |  |  |  |
| Early breast cancer versus advanced or metastatic breast cancer: *p<0.05, **p<0.01, ***p<0.001 |  |  |  |  |
| HER2E: HER2 enriched, TNBC: tripple negative breast cancer |  |  |  |  |

### Independent variables associated with advanced diagnosis of BC

There was no mean difference in age at diagnosis between patients presented with early disease and those with advanced disease (52.30 vs. 51.23, p=0.056). There was no mean difference in BMI between patients diagnosed at early stages and those diagnosed at advanced stages (31.08 vs. 31.66, p=0.219). Positive family history was significantly more common among patients who presented at early stages, compared to those who presented at advanced stages (33.5% vs. 28.4%, p=0.016). Having paid job was less common among patients with advanced disease, compared to patients with early disease (40.1% vs. 46.1%, p=0.009). Percentage of unmarried patients was higher in the early-stage group than in advanced-stage group (25.8% vs. 21.5%, p=0.031).

Regarding residence, the percentage of rural residents was higher in the advanced-stage group than in early-stage group (9.1% vs. 5.0%, p=0.001). Percentage of Cairo and Giza residents was higher in the early-stage group than the advanced-stage group (66.0% vs. 60.0%, p=0.012). Figure 1 shows the ascending ranking of advanced cases’ percentages in each city of Egypt. El Wadi El Gedeed city had the highest rank, while El Bahr El Ahmar had the lowest rank. The figure also shows that some cities of Upper Egypt that are in the south (Luxor, Aswan, Minya, Qena, and Sohag) got the highest ranking compared to the cities that are in the north and contain the Nile Delta. There was no statistically significant association between religion and stage at diagnosis (p=0.398).

**Figure 1.**
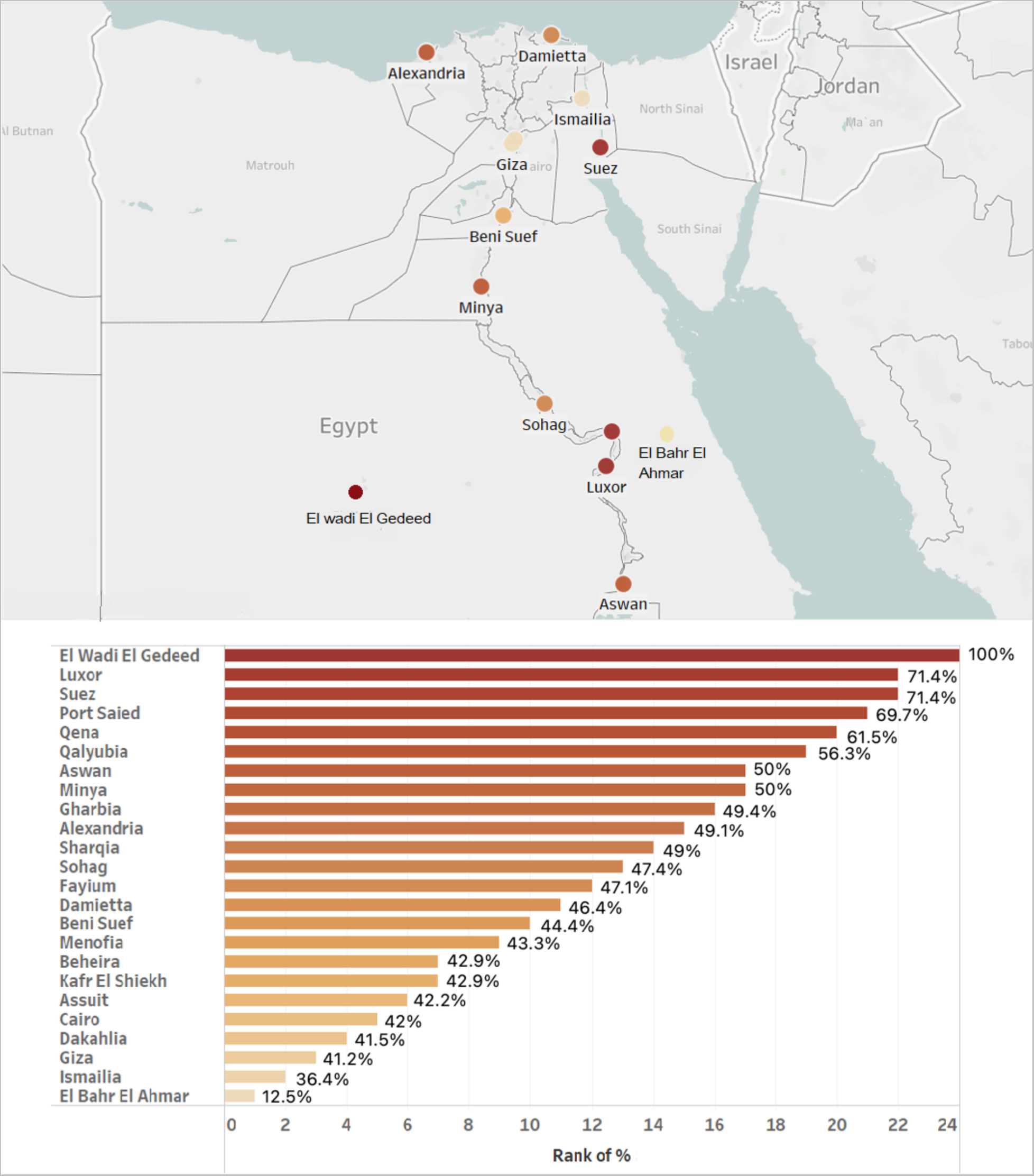
Ascending ranking of advanced cases’ percentages by geographical location

Having more than three children was more common in the advanced-stage group compared to early-stage (28.9% vs. 22.4%, p=0.001). There was no statistically significant association between gender and stage at diagnosis (p=0.204). Percentage of patients with HER2 or TNBC subtypes was higher in the advanced-stage group compared to early-stage group (29.2% vs. 23.2%, p=0.005). The mean difference in KI-67% score was 2.79 favoring the advanced disease group (p<0.001). Percentage of patients with tumor grade I was higher in the early-stage group compared to advanced-stage group (2.9% vs. 0.7%, p=0.007). There was no statistically significant association between menopausal status and stage at diagnosis (p=0.213) (Table 1).

### Univariate regression analysis of the association between the independent variables and advanced stage diagnosis of BC

Table 2 shows the unadjusted odds ratios (ORs) with 95% confidence interval (CI) of getting diagnosed at advanced stages. One additional year increase in age was associated with 1% decrease in the odds of presenting with advanced disease, but it was not statistically significant (OR=0.99; 95% CI: 0.99 - 1.00, p=0.059). Patients with negative family history of BC had 27% higher odds of presenting with advanced disease compared to patients with positive family history (OR= 1.27; 95% CI: 1.05 – 1.54, p=0.016). Patients with no job had 28% higher odds of presenting with advanced disease compared to patients with paid job (OR= 1.28; 95% CI: 1.06 – 1.53, p=0.009). Married patients were 27% more likely to present at advanced stages compared to unmarried patients (OR=1.27; 95% CI: 1.02 – 1.57, p=0.031).

**Table 2.** Unadjusted and adjusted OR with 95% CI of getting diagnosed at advanced or metastatic stages.

|  | Advanced or metastatic stage diagnosis of breast cancer<br>n= 855 |  |  |  |
| --- | --- | --- | --- | --- |
|  | Unadjusted OR (95% CI) | p-value | Adjusted OR (95% CI) | p-value |
| **Sociodemographic variables** |  |  |  |  |
| Age (years) | 0.99 (0.99 - 1.00) | 0.059 | 1.00 (0.99 - 1.01) | 0.956 |
| Body mass index (kg/m2) | 1.01 (0.99 - 1.03) | 0.231 | 1.01 (0.99 - 1.04) | 0.325 |
| KI-67% score | **1.01 (1.00 - 1.01)\*** | 0.022 | **1.01 (1.00 - 1.02)\*** | 0.013 |
| Family history of BC |  |  |  |  |
| Negative family history | **1.27 (1.05 – 1.54)\*** | 0.016 | 1.00 (0.76 - 1.32) | 0.986 |
| Positive family history | Reference |  | Reference |  |
| Employment status |  |  |  |  |
| No job (housewife, retired, or student) | **1.28 (1.06 - 1.53)\*\*** | 0.009 | **1.44 (1.10 - 1.90)\*\*** | 0.009 |
| In paid job | Reference |  | Reference |  |
| Marital status |  |  |  |  |
| Married | **1.27 (1.02 – 1.57)\*** | 0.031 | 1.18 (0.87 – 1.60) | 0.291 |
| Unmarried (single, divorced, or widow/widower) | Reference |  | Reference |  |
| Urbanicity |  |  |  |  |
| Rural | **1.89 (1.31 - 2.72)\*\*** | 0.001 | **1.88 (1.03 - 3.42)\*** | 0.040 |
| Urban | Reference |  | Reference |  |
| City of residence |  |  |  |  |
| Other 22 cities | **1.30 (1.08 – 1.56)\*\*** | 0.007 | 0.96 (0.71 - 1.78) | 0.785 |
| Big cities (Cairo and Giza) | Reference |  | Reference |  |
| Religion |  |  |  |  |
| Christian | 1.18 (0.80 - 1.73) | 0.399 | 1.44 (0.85 - 2.45) | 0.180 |
| Muslim | Reference |  | Reference |  |
| Parity |  |  |  |  |
| More than 3 children | **1.42 (1.15 – 1.75)\*\*** | 0.001 | 1.30 (0.94 – 1.78) | 0.113 |
| 3 children or fewer | Reference |  | Reference |  |

| Advanced or metastatic stage diagnosis of breast cancer |  |  |  |  |
| --- | --- | --- | --- | --- |
| n= 855 |  |  |  |  |
|  | Unadjusted OR (95% CI) | p-value | Adjusted OR (95% CI) | p-value |
| Gender |  |  |  |  |
| Male | 1.94 (0.69 – 5.46) | 0.212 | 2.07 (0.45 – 9.63) | 0.352 |
| Female | Reference |  | Reference |  |
| **Biological variables** |  |  |  |  |
| Breast cancer subtypes |  |  |  |  |
| HER2E or TNBC | **1.44 (1.16 - 1.79)\*\*** | 0.001 | 0.93 (0.66 - 1.31) | 0.667 |
| Other subtypes (Luminal A, B1 and B2) | Reference |  | Reference |  |
| Tumor grade |  |  |  |  |
| Grade II | **4.12 (1.58 – 10.77)\*\*** | 0.004 | 1.51 (0.47 – 4.83) | 0.488 |
| Grade III | **4.35 (1.62 – 11.70)\*\*** | 0.004 | 1.42 (0.42 – 4.86) | 0.575 |
| Grade I | Reference |  | Reference |  |
| Menopausal status |  |  |  |  |
| Premenopausal | 1.13 (0.93 – 1.36) | 0.214 | 1.08 (0.80 – 1.44) | 0.629 |
| Postmenopausal | Reference |  | Reference |  |
| Bold indicates statistically significant findings by univariate and multivariate logistic regression<br>OR of being diagnosed at advanced stages compared to early stages: *p<0.05, **p<0.01, ***p<0.001<br>HER2E: HER2 enriched, TNBC: tripple negative breast cancer<br>All variables were adjusted for family history, employment status, marital status, urbanicity, city of residence, parity, breast cancer subtype, tumor grade, and KI-67% score (significant variables in the univariate logistic regression analysis) |  |  |  |  |

Rural residents were 89% more likely to present with advanced disease compared to urban residents (OR=1.89; 95% CI: 1.31 – 2.72, p=0.001). Patients who reside in distant cities had 30% higher odds of getting diagnosed at advanced stages compared to those who reside in Cairo or its urban extension Giza (OR= 1.30; 95% CI: 1.08 – 1.56). There was no statistically significant difference in odds of presenting with advanced disease between Christian and Muslim patients (OR=1.18; 95% CI= 0.80 – 1.73, p=0.399). Having more than three children was associated with 42% higher odds compared to having three children or fewer (OR=1.42; 95% CI: 1.15– 1.75, p=0.001). There was no statistically significant difference in odds of getting diagnosed at advanced stages between females and males (OR=1.94; 95% CI: 0.69 – 5.46, p=0.212).

Patients with HER2E or TNBC subtypes were 44% more likely to get diagnosed at advanced stages compared to patients with other subtypes (OR= 1.44; 95% CI: 1.16 – 1.79, p=0.001). One additional unit increase in KI-67% score was significantly associated with 1% increase in the odds of presenting with advanced disease (OR=1.01; 95% CI: 1.00 1.01, p=0.022). Patients with tumor grade III had higher odds of presenting with advanced disease compared to patients with tumor grade I (OR=4.35; 95% CI: 1.62 – 11.70, p=0.004). Patients with tumor grade II had higher odds of presenting with advanced disease compared to patients with tumor grade I (OR=4.12; 95% CI: 1.58 – 10.77, p=0.004). There was no statistically significant difference of presenting at advanced stages between postmenopausal and premenopausal women (OR=1.13; 95% CI: 0.93 – 1.36, p=0.214). Figure 2 shows a forest plot for unadjusted ORs with 95% CI of getting diagnosed at advanced stages.

**Figure 2.**
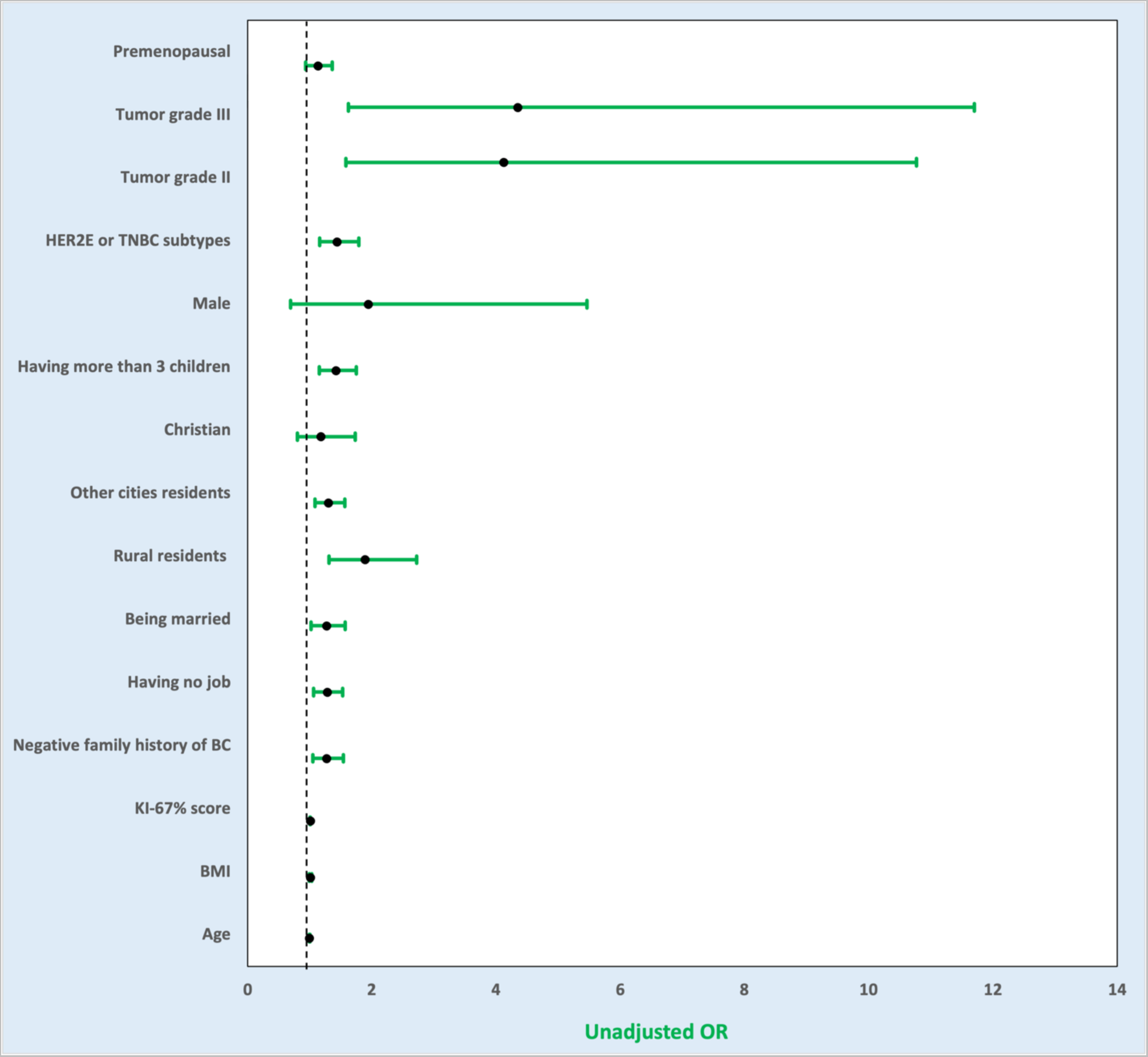
Forest plot for unadjusted ORs of advanced stage diagnosis of breast cancer

### Multivariate regression analysis of the association between the independent variables and advanced stage diagnosis of BC

All variables were adjusted for KI-67% score, employment status, marital status, urbanicity, city of residence, parity, BC subtype, and tumor grade. Only KI-67% score, employment status, and urbanicity were statistically significantly associated with advanced-stage diagnosis of BC. One additional unit increase in KI-67% score was associated with 1% increase in the odds of presenting with advanced disease (aOR=1.01; 95% CI: 1.00 1.02, p=0.013). Patients with no job had 44% higher odds of presenting with advanced disease compared to patients with paid job (aOR=1.44; 95% CI: 1.10 – 1.90, p=0.009). Patients reside in rural areas had 88% higher odds of presenting with advanced disease compared to patients reside in urban areas (aOR=1.88; 95% CI: 1.03 – 3.42, p=0.040) (Table 2). Figure 3 shows a forest plot for adjusted ORs with 95% CI of getting diagnosed at advanced stages.

**Figure 3.**
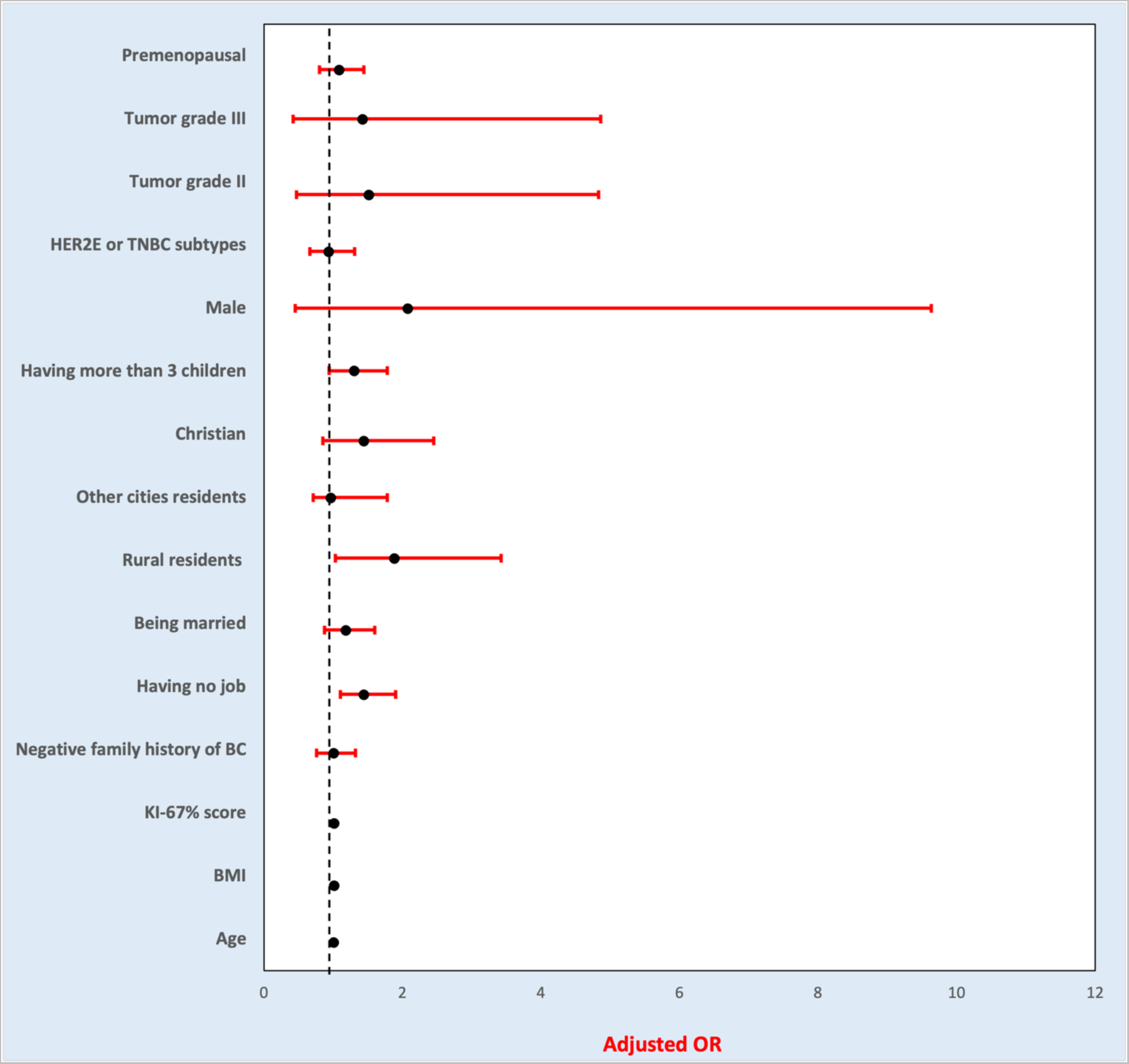
Forest plot for adjusted ORs of advanced stage diagnosis of breast cancer

## Discussion

This study shed light on the sociodemographic disparities associated with late presentation of BC in Egypt. In the univariate models, rural residency, being married, having no job, having negative family history of BC, having more than three children, having TNBC or HER2E BC subtypes, having high tumor grade, and having high KI-67% score were associated with advanced disease. In the multivariate model, only kI-67% score, rural residency, and having no job were significantly associated with advanced stage diagnosis of BC. Age, menopausal status, BMI, gender, and religion were not associated with advanced disease, neither in the univariate nor multivariate regression models. To our knowledge, this is the first study to explore the impact of these inequalities among Egyptian BC patients.

Previous studies from Iran, New Zealand, Nigeria, and North-west Ethiopia observed a significant association between urbanicity and BC stage at diagnosis. Rural residents had higher odds of presenting with late-stage BC compared to urban residents (7*–*10). This finding is consistent with this cohort’s finding. The association between rural residency and advanced diagnosis of BC can be explained through different indirect relationships. Due to lack of awareness campaigns and presence of limited number of specialized oncology facilities in rural areas in Egypt, rural residents are less likely to utilize breast cancer screening and early detection techniques which can lead to late diagnosis of BC. Rural residents and especially women living in rural areas are less likely to be offered well education and good paying jobs. Therefore, many of them cannot afford neither acess to screening facilities nor traveling to big cities to undergo the needed imaging, biopsies, etc.

Previous studies claimed that unmarried patients were more likely to get diagnosed at advanced stages compared to married patients (4,11). Interestingly, unmarried women were less likely to present with advanced disease in this study. This finding is not consistent with majority of previous studies, except one study that has been conducted in Pakistan which claimed that unmarried patients had shorter delay in seeking medical help compared to married patients (12). The association between being married and advanced-stage diagnosis of BC can be explained through few indirect relationships. Fear of separation, heavy marital responsibilities, and financial struggles could delay married people from seeking medical help earlier.

The association between family history and BC stage at diagnosis were controversial in the previous studies [10,50]. In this study, patients with negative family history of BC were more likely to get diagnosed late compared to patients with positive family history. This can be explained by that people with negative family history of BC are less aware of BC symptoms and its screening techniques which would make them present at late stages.

Majority of previous studies demonstrated that younger patients were more likely to get diagnosed at advanced stages compared to older patients (8,11,13–18). In contrast, this cohort observed no significant association between younger age and advanced stage diagnosis of BC in Egypt, which can be explained by lack of BC awareness and lack or breast cancer screening utilization among all age groups in Egypt.

Besides the sociodemographic disparities that are linked to advanced-stage diagnosis of BC, there are some clinical factors that are linked to advanced-stage diagnosis of BC as well. KI-67% score was significantly associated with BC stage at diagnosis in this cohort. Nevertheless, it was the major confounder that confounded the association between stage at diagnosis and other variables that turned to be nonsignificant in the multivariate regression analysis.

The findings of this cohort revealed the catastrophic state of the health sector in Egypt. Public hospitals in Egypt are from bad to worse; there are many challenges such as lack of accountability, uncontrolled corruption, underfunding, lack of medical equipment, low quality of medical care, lack of qualified personnel (19,20). Private hospitals have unaffordable medical care which results in deprivation and even death of many patients (19). This can be addressed by increasing the expenditure on health, reforming the health system’s organizational structure and management, increasing the healthcare professionals’ salaries, and providing public hospitals with adequate medical equipment (21). Since early detection of breast cancer is the key point towards good prognosis and longer survival rate, raising the population awareness on breast cancer symptoms and breast cancer screening techniques is mandatory. Raising the population awareness on different public health issues is the best approach to decrease the financial and medical burden on the families, hospitals, and the government. Moreover, raising awareness is considered as one of the less stigmatized approaches in promoting population health (22). This can be achieved through implementing health campaigns in deprived and rural areas and implementing advertising campaigns on different platforms such as television, radio, and social media.

This study is the first to be conducted in Egypt. The study included many sociodemographic variables that were underexamined in the literature such as gender, religion, and BMI. The study also considered the clinical factors while examining the sociodemographic factors such as BC subtypes, tumor grade, and KI-67% score; to eliminate any possible confounding impact of the clinical factors on the association between BC stage at diagnosis and sociodemographic factors.

This cohort study is limited in terms of generalizability because data were collected from one single center in Egypt. Therefore, conducting this study on a larger scale is recommended. Since data were collected retrospectively from the medical files, there were many individuals whose clinical and sociodemographic data were missing. In view of that, this study excluded the individuals with incomplete data; exclusion of individuals with incomplete data could potentially contribute to selection bias. Additionally, patients’ religion was interpreted based on their names which could be biased and accordingly being agnostic was not taken into consideration in this cohort. Further research could examine other sociodemographic variables such as level of education, monthly income, socioeconomic status, and distance to medical facilities. Furthermore, conducting interviews with BC patients who have advanced disease would help recognize the reasons behind their delay in seeking medical help then accordingly guide policy towards better outcomes.

## Conclusion

This 10-year single-center study highlights the sociodemographic disparities associated with advanced and late-stage diagnosis of BC in Egypt. Higher kI-67% scores, rural residence, and having no job were significantly associated with advanced stage diagnosis of BC. With the recent implementation of a nationwide screening program in Egypt, specific measures should be taken to address the advanced-stage diagnosis in these relevant subgroups. Adjusting expenditure on healthcare and education to address similar public health challenges will help promote the health and economic status of Egyptians. In addition, raising the physician and population awareness is the best strategy to encourage early detection of BC. Finally, supporting public health research in Egypt would help understand more the sociodemographic inequalities among patients to improve the health outcomes and life expectancy of Egyptians.

## Data Availability

The data used in this cohort are available upon request from the corresponding author.

## Abbreviations

BC: breast cancer
HER2E: HER2 enriched
TNBC: triple-negative breast cancer;
BMI: body mass index

## Authors’ contributions

MAF, HAA, LK, and JCY contributed to developing study concept and design.

MAF, HAA,LK, and JCY contributed to data analysis and interpretation.

MAF, HAA, LK, and JCY contributed to writing the initial draft of the manuscript.

JCY, LK, and HAA contributed to reviewing the whole study.

All authors approved the final version of the manuscript.

## Availability of data and materials

The data used in this cohort are available upon request from the corresponding author.

## Ethical approval and informed consent of participants

The study was approved by the local IRB (Research Ethics Committee of Cairo Oncology Center) in November 2022 under a case reference number of COC-2022013.

## Supplementary Figures

## References

1. WHO. Breast cancer. 2021 Mar; Available from: https://www.who.int/news-room/fact-sheets/detail/breast-cancer

2. Ibrahim AH, Shash E. General Oncology Care in Egypt. In: Al-Shamsi HO, Abu-Gheida IH, Iqbal F, Al-Awadhi A, editors. Cancer in the Arab World [Internet]. Singapore: Springer Singapore; 2022 [cited 2023 Mar 6]. p. 41–61. Available from: https://link.springer.com/10.1007/978-981-16-7945-2_4

3. Azim HA, Elghazawy H, Ghazy RM, Abdelaziz AH, Abdelsalam M, Elzorkany A, et al. Clinicopathologic Features of Breast Cancer in Egypt—Contemporary Profile and Future Needs: A Systematic Review and Meta-Analysis. JCO Glob Oncol [Internet]. 2023 Mar [cited 2023 Jul 24];(9):e2200387. Available from: https://ascopubs.org/doi/10.1200/GO.22.00387

4. dos-Santos-Silva I, De Stavola BL, Renna NL, Nogueira MC, Aquino EML, Bustamante-Teixeira MT, et al. Ethnoracial and social trends in breast cancer staging at diagnosis in Brazil, 2001–14: a case only analysis. Lancet Glob Health [Internet]. 2019 Jun [cited 2023 Mar 6];7(6):e784–97. Available from: https://linkinghub.elsevier.com/retrieve/pii/S2214109X19301512

5. Amr Mohamed Kandil. Sisi proposes creating fund to tackle women’s challenges. Egypt Today [Internet]. 2019 Mar; Available from: https://www.egypttoday.com/Article/1/67718/Sisi-proposes-creating-fund-to-tackle-women-s-challenges

6. Taheri M, Tavakol M, Akbari ME, Almasi-Hashiani A, Abbasi M. Relationship of Socio Economic Status, Income, and Education with the Survival Rate of Breast Cancer: A Meta-Analysis. Iran J Public Health. 2019 Aug;48(8):1428*–*38.

7. Foroozani E, Ghiasvand R, Mohammadianpanah M, Afrashteh S, Bastam D, Kashefi F, et al. Determinants of delay in diagnosis and end stage at presentation among breast cancer patients in Iran: a multi-center study. Sci Rep [Internet]. 2020 Dec 8 [cited 2023 Mar 6];10(1):21477. Available from: https://www.nature.com/articles/s41598-020-78517-6

8. Seneviratne S, Lawrenson R, Harvey V, Ramsaroop R, Elwood M, Scott N, et al. Stage of breast cancer at diagnosis in New Zealand: impacts of socio-demographic factors, breast cancer screening and biology. BMC Cancer [Internet]. 2016 Dec [cited 2023 Mar 6];16(1):129. Available from: http://www.biomedcentral.com/1471-2407/16/129

9. Jedy-Agba E, McCormack V, Olaomi O, Badejo W, Yilkudi M, Yawe T, et al. Determinants of stage at diagnosis of breast cancer in Nigerian women: sociodemographic, breast cancer awareness, health care access and clinical factors. Cancer Causes Control [Internet]. 2017 Jul [cited 2023 Mar 6];28(7):685–97. Available from: http://link.springer.com/10.1007/s10552-017-0894-y

10. Tesfaw A, Tiruneh M, Tamire T, Yosef T. Factors associated with advanced stage diagnosis of breast cancer in North West Ethiopia. a cross-sectional study. ecancermedicalscience [Internet]. 2021 Mar 25 [cited 2023 Mar 6];15. Available from: https://ecancer.org/en/journal/article/1214-factors-associated-with-advanced-stage-diagnosis-of-breast-cancer-in-north-west-ethiopia-a-cross-sectional-study

11. Koroukian SM, Bakaki PM, Htoo PT, Han X, Schluchter M, Owusu C, et al. The Breast and Cervical Cancer Early Detection Program, Medicaid, and breast cancer outcomes among Ohio’s underserved women: Disparities in Breast Cancer Outcomes. Cancer [Internet]. 2017 Aug 15 [cited 2023 Mar 6];123(16):3097–106. Available from: https://onlinelibrary.wiley.com/doi/10.1002/cncr.30720

12. Majeed I, Ammanuallah R, Anwa AW, Rafique HM, Imran F. Diagnostic and treatment delays in breast cancer in association with multiple factors in Pakistan. East Mediterr Health J [Internet]. 2021 Jan 1 [cited 2023 Mar 6];27(1):23–32. Available from: https://applications.emro.who.int/emhj/v27/01/1020-3397-2021-2701-23-32-eng.pdf

13. Agodirin O, Aremu I, Rahman G, Olatoke S, Olaogun J, Akande H, et al. Determinants of Delayed Presentation and Advanced-Stage Diagnosis of Breast Cancer in Africa: A Systematic Review and Meta-Analysis. Asian Pac J Cancer Prev [Internet]. 2021 Apr 1 [cited 2023 Mar 6];22(4):1007–17. Available from: http://journal.waocp.org/article_89562.html

14. de Oliveira NPD, de Camargo Cancela M, Martins LFL, de Souza DLB. A multilevel assessment of the social determinants associated with the late stage diagnosis of breast cancer. Sci Rep [Internet]. 2021 Feb 1 [cited 2023 Mar 6];11(1):2712. Available from: https://www.nature.com/articles/s41598-021-82047-0

15. Sealy-Jefferson S, Roseland ME, Cote ML, Lehman A, Whitsel EA, Mustafaa FN, et al. Rural– Urban Residence and Stage at Breast Cancer Diagnosis Among Postmenopausal Women: The Women’s Health Initiative. J Womens Health [Internet]. 2019 Feb [cited 2023 Mar 6];28(2):276–83. Available from: https://www.liebertpub.com/doi/10.1089/jwh.2017.6884

16. DeGuzman PB, Cohn WF, Camacho F, Edwards BL, Sturz VN, Schroen AT. Impact of Urban Neighborhood Disadvantage on Late Stage Breast Cancer Diagnosis in Virginia. J Urban Health [Internet]. 2017 Apr [cited 2023 Mar 6];94(2):199–210. Available from: http://link.springer.com/10.1007/s11524-017-0142-5

17. Franzoi MA, Rosa DD, Zaffaroni F, Werutsky G, Simon S, Bines J, et al. Advanced Stage at Diagnosis and Worse Clinicopathologic Features in Young Women with Breast Cancer in Brazil: A Subanalysis of the AMAZONA III Study (GBECAM 0115). J Glob Oncol [Internet]. 2019 Dec [cited 2023 Mar 6];(5):1–10. Available from: https://ascopubs.org/doi/10.1200/JGO.19.00263

18. Miller JW, Royalty J, Henley J, White A, Richardson LC. Breast and cervical cancers diagnosed and stage at diagnosis among women served through the National Breast and Cervical Cancer Early Detection Program. Cancer Causes Control [Internet]. 2015 May [cited 2023 Mar 6];26(5):741–7. Available from: http://link.springer.com/10.1007/s10552-015-0543-2

19. Soha Bayoumi. Health and social justice in Egypt: towards a health equity perspective. In 2016. Available from: https://en.unesco.org/inclusivepolicylab/sites/default/files/analytics/document/2019/4/wssr_2016_chap_30.pdf

20. EGYPT | Summary. Available from: https://www.publichealth.columbia.edu/research/others/comparative-health-policy-library/egypt-summary#:~:text=The%20public%20health%20system%20faces,medical%20equipment%2C%20and%20qualified%20personnel.&text=Government%20investment%20in%20the%20public,GDP%20for%20public%20health%20expenditures.

21. Nesreen M. Kamal Elden, Hoda I. Ibrahim Rizk, Ghada Wahby. Improving Health System in Egypt: Perspectives of Physicians. Egypt J Community Med [Internet]. 2016 Jan 1 [cited 2023 Mar 31];34(1):45–58. Available from: http://ejcm.journals.ekb.eg/article_646.html

22. Tsai CL, Lin YW, Hsu HC, Lou ML, Lane HY, Tu CH, et al. Effects of the Health-Awareness-Strengthening Lifestyle Program in a Randomized Trial of Young Adults with an At-Risk Mental State. Int J Environ Res Public Health [Internet]. 2021 Feb 18 [cited 2023 Mar 31];18(4):1959. Available from: https://www.mdpi.com/1660-4601/18/4/1959

